# Association of indoor use of pesticides with CKD of unknown origin

**DOI:** 10.1101/2022.10.21.22281385

**Authors:** Saba Alvand, Sudabeh Alatab, Sahar Dalvand, Fariba Shahraki-Sanavi, Mahmoud Ali Kaykhaei, Elham Shahraki, Erfaneh Barar, Sadaf G Sepanlou, Alireza Ansari-Moghaddam

## Abstract

Diabetes mellitus and hypertension are the two main etiologies of chronic kidney disease (CKD). However, CKD subjects of unknown origin (CKDu) have been recognized recently. One of the proposed causes is pesticide use in farmers. On the other hand, house use of pesticides has never been investigated in developing countries. In this study, we aimed to investigate the association between house use of pesticide and their exposure time with CKDu. This study is part of the population-based cohort of Prospective Epidemiological Research Studies in Iran. We used the baseline data of the Zahedan Adult Cohort Study. We dropped all subjects with diabetes mellitus and/ or hypertension, egfr 60-89 ml/min/1.73 m^2^, and unavailable creatinine measurement. Subjects with egfr of less than 60 ml/min/1.73 m^2^ through MDRD equations were defined as CKDu and compared with subjects with egfr of more than 90 ml/min/1.73 m^2^. The house use of pesticides and exposure time were asked through a questionnaire. In this study 10072 participants enrolled, and 1079 remained in the final sample after appliance the exclusion criteria. Female sex, single marital status, low physical activity, TG more than 150 mg/dl, BMI of more than 25, non-smokers, house use of pesticide, and more time exposed to pesticides were associated with CKDu. The effects of age, female sex, TG more than 150 mg/dl, pesticide use (1.36;95%CI 1.01-1.84), and the third tertile of exposure time compared to non-users (1.64;95%CI 1.07-2.51) remained significant in multivariable analysis. We found a positive association between the use and exposure time with kidney function in cases without diabetes mellitus and hypertension. Further longitudinal studies should be carried out to assess this effect.

## Introduction

Chronic kidney disease (CKD) as a non-communicable disease has placed a high economic burden on communities to provide high-cost therapies, kidney transplantation, and dialysis, for patients at end stage renal disease. Moreover, CKD is a recognized independent risk factor for cardiovascular disease events, which is the main reason for death in these patients(1). Based on the most recent analysis of the Global Burden of Disease study 2017, it is estimated that more than 700 million individuals worldwide are affected by CKD. This increase is more prominent in lower and middle-income economically developing countries including countries in North Africa and Middle East(2). Accordingly, a meta-analysis study in Iran reported the overall prevalence of CKD at 15.14% with a high variation among provinces, a figure that indicating a higher than the global average prevalence of CKD in our country(3).

Diabetes mellitus type 2 (T2DM) and hypertension (HTN) are the two main etiologies of CKD. Up to 34% of CKD dalys are due to diabetes mellitus alone (2). However, in the last 20 years, a new term “CKD of unknown origin (CKDu)” has been introduced in areas of Central America and South Asia that address the diagnosis of CKD which is not attributable to any traditional risk factor, such as diabetes mellitus, hypertension, glomerular nephritis, obstructive nephropathy or congenital structural abnormalities. The term CKDu was first used in El Salvador in the early 2000s to describe a disease predominantly affecting agricultural communities. From then, the CKDu is being reported with increasing frequency from predominantly rural locations in several regions across the world(4).

Many hypotheses have been proposed for assessment of potential causes of CKDu. Chronic dehydration and agrochemicals (pesticides, herbicides, fertilizers) use were the most frequent ones assessed in several studies because CKDu is mainly seen in rural men with agricultural occupation(5–7). However other environmental factors, such as heavy metal exposures, high seasonal temperatures, mycotoxins, contaminated water supplies, and snake bite, also being considered(8–11).

Data regarding prevalence and epidemiological features of CKDu is scanty. In our country, we have Epidemiological Research Studies in iran (PERSIAN) which is a nationwide cohort study that collects data from participants 35-70 years from 18 cohort centers in 16 provinces in Iran(12). Zahedan cohort study (ZACS) is part of the PERSIAN cohort that has been carried out in Zahedan, the capital city of Sistan and Baluchestan. Sistan and Baluchestan is exceptional area compared to other provinces due to the scarcity of population, climate, ethnicity, lifestyle, and the least favorable health status (13). Although agricultural occupations are scarce in this region, pesticides use in yards and homes are far more common than in other parts of Iran due to high number of vermin (14). Thus, Zahedan is a proper location to investigate the prevalence of CKDu and assess its possible determinants including indoor use of pesticides. Therefore, the purpose of this study was to estimate the prevalence of low egfr (< 60 ml/min /1.7 m^2^, calculated by MDRD equation) in those without known HTN, T2DM in this particular region and also to assess its main contributors including house use of pesticide and their exposure time among participants.

## Materials and methods

### Study design & location

This study was a cross-sectional analysis of data from ZACS as part of PERSIAN project. The detailed explanation of PERSIAN study and zacs has been published previously(12,15). In brief, from October 2015 to January 2019, 10,079 participants who aged 35-70 years and resided for at least nine months in these regions (based on the regional records) enrolled in zacs study. Participants with physical or psychological disabilities who could not complete the process were excluded.

Written informed consent was obtained from all participants before enrollment. This study was approved by the ethics committee Zahedan University of Medical Sciences as the main executive university with the approval number of IR.ZAUMS.REC.1401.160.

Sistan and Baluchestan, the second largest province in Iran, is located in the southeast of Iran, neighboring Pakistan and Afghanistan, with 2.7 million people reside in a 180 726 km2 area. The lowest human development index (HDI) in Iran in 2014 belonged to Sistan and Baluchestan province, with an index of 0.589 out of the 31 provinces(16). The population density is 15 persons per square km, much lower than the average population density in Iran, which is 52 persons per square km. This province is one of the driest regions in the country and is categorized as a hot desert climate. However, the coastal areas have a humid climate due to the border with the Gulf of Oman. Current study took part in three districts of Sistan and Baluchestan with low, middle, and high socioeconomic status. These three districts were chosen randomly based on having the major ethnic groups, large geographic area, and low migration rate(15).

The data were collected by employing questionnaires through interviews. The PERSIAN cohort questionnaires consist of three parts: general, medical, and nutrition, and each part contain different topics. The complete data dictionary is available on the PERSIAN cohort site (https:/persiancohort.com/). Personal information, anthropometry, physical activity, house use of pesticide and duration of exposure were collected from the general section. Past medical history, complete list of medications, and personal habits, were gathered from the medical section. The anthropometrics were obtained in the morning after collecting samples when the participants were still fasting. All the centers used similar adjusted tools for the measurements. Weight was measured by Seca 762 mechanical flat scale in kilograms, and other anthropometrics were measured by Seca 206 body meter measuring tape in centimeters. The trained personnel measured blood pressure with Riester auscultatory sphygmomanometers based on the American heart association criteria(12,17).

Blood sample was collected from all participants after 8–12 h of fasting. The blood samples were transferred to the PERSIAN cohort reference laboratory within 3 h of sampling for measuring complete blood count, fasting blood sugar (FBS), total cholesterol, high-density lipoprotein cholesterol (HDL), triglycerides (TG), alanine transaminase, aspartate transaminase, alkaline phosphatase, gamma-glutamyl transpeptidase, blood urea nitrogen, and creatinine levels.

### Definitions

HTN was defined as having any of the following conditions: self-report use of hypertension medications, anti-hypertensive medication consumption, systolic blood pressure ≥ 140 mmhg, or diastolic blood pressure (DBP) ≥ 90 mmhg (18).

Diabetes Mellitus was defined as having any of the following conditions : self-report use of diabetes mellitus medications, blood glucose-lowering medications consumption, FBS≥126 mg/dl (19). People with FBS< 126 mg/dl were considered healthy in this study.

Physical activity status included in the analysis as tertiles by metabolic equivalent task (MET) score (20).

Pesticide exposure was evaluated based on the detailed questionnaire asking about the use, frequency and duration of exposure to pesticide(s). In case of a positive exposure (yes/ no answer), the number and duration of exposure were asked. The sum of each exposure duration was calculated and expressed in minutes. The duration of pesticide exposures in lifetime was treated as a continuous variable and included in the analysis in three groups of less than 9 minutes, between 9-20 minutes and more than 20 minutes based on tertiles.

The standard colorimetric Jaffe-Kinetic reaction method were used to measure serum creatinine levels, which was not traceable to isotope dilution mass spectroscopy (IDMS). Since we did not have access to albuminuria, the kidney damage was investigated by GFR measurement alone. MDRD study equation was applied for estimating GFR based on the following formula:

GFR by MDRD (ml/min/1.73 m^2^) = 176 × Cr^−1.154^ × age^−0.203^ × 0.742 (if female)

CKD was based on one measurement of serum creatinine and defined as participants who had egfr less than 60 ml/min/1.73 m^2^ (21,22). Participants with egfr higher than 90 ml/min/1.73 m^2^ were counted as normal egfr due to unclear kidney damage in this spectrum without urine data(23).

CKDu group defined as participants who had egfr less than 60 ml/min/1.73 m^2^ without diabetes mellitus and HTN (based on the mentioned definitions). Non-CKDu group defined as subjects with egfr higher than 90 ml/min/1.73 m^2^ without diabetes and HTN. Since this study is nested from a large population-based cohort, exclusion of other known etiologies for CKDu through ultra-sonography (to exclude structural kidney diseases) and urine analysis (to exclude glomerular diseases) were not cost-benefit and applicable.

### Statistical analysis

Frequency and percentage were used to describe qualitative variables, and a comparison of CKDu and non-CKDu groups was assessed through a Chi-squared test. Physical activity and house use of pesticide exposure time were grouped based on their tertiles. TG and HDL were used as binary variables with a cut-off of equal or more than 150 mg/dl for TG (named hyper TG) and less than 40 mg/dl for males and 50mg/dl for females in HDL (named low HDL).

Mean and standard deviation were used to describe quantitative variables, and a student t-test was applied to check the difference between the two main groups of the study.

In this study, crude odds ratios (OR) and their 95% confidence intervals (95% CI) were calculated using univariable logistic regression to explore the unadjusted association between variables and outcome. Variables with p-values smaller than 0.20 were included for further multivariable analysis. Accordingly, we used of two multivariable logistic regression models to evaluate the relationship between CKDu with age, sex, marital status, BMI, smoking, hyper TG and house use of pesticides in model 1; and CKDu with the age, sex, marital status, BMI, smoking, hyper TG and house use of pesticides exposure time in model 2. The statistical significance was declared if the p-value was less than 0.05, and all the analyses were performed using STATA (version 12; statacorp).

## Results

During the study period, 10,072 participants enrolled in the ZACS from which 9,977 subjects had blood samples and therefore included in the study. Complete flow diagram of study is presented in Fig 1. The mean age of participants was 50.4 years (SD= 9.2), and 39.1% were male. The prevalence of CKD in our sample was 9.1 % (95% CI: 8.6-9.6) which was 5.0% (95% CI: 4.4-5.7) in males and 13.3% in females (95% CI: 12.4-14.1).

**Fig 1.**
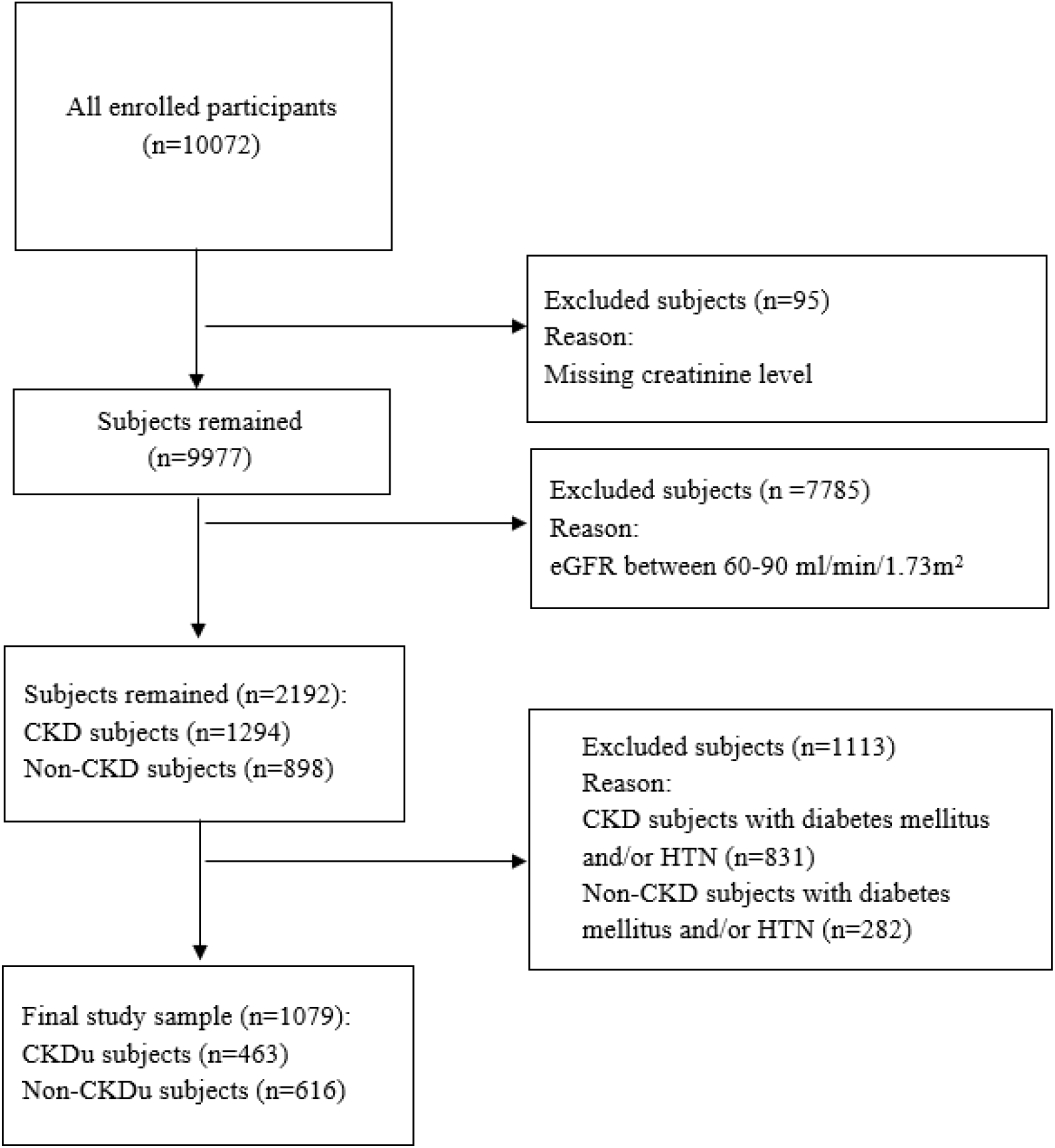
Study sample flow diagram.

In our population, 7,785 (78%) subjects had egfr between 60-89 ml/min/1.73m^2^ and therefore excluded from our analysis. Among remaining 2,192 subjects, the prevalence of CKD was 59.03% (egfr less than 60 ml/min/1.73m^2^, n= 1294). The prevalence of subjects with diabetes mellitus and/ or HTN was 64.2% in our CKD group and 31.4% in non-CKD group (Fig 1).

Among the 1,079 remaining subjects, 463 subjects (42.9%) had egfr less than 60 ml/min/1.73m^2^ (CKDu group) and 616 participants had egfr higher than 90 ml/min/1.73m^2^ (non-CKDu group). All CKDu subjects were in stage 3.

The mean age of subjects in CKDu group was significantly higher than non-CKDu group (52.1 ± 9.1 years vs. 45.6 ± 8.2; p=0.02). The male subjects comprise 61.4% of non-CKDu while in CKDu this percentage was significantly lower and reached 20.7% (p<0.001). The frequency of no use of pesticides was significantly higher in non-CKDu compared to CKD-u (59.3% vs 49.7%, p= 0.002). Accordingly, higher exposed time to pesticides was mainly seen in subjects with CKDu (p= 0.01). Table 1 demonstrates the details of the demographics and medical history of the included subjects in two groups.

**Table 1.** Demographics, medical history, and house use of pesticides in the studied population.

| Parameters |  | Non-CKDu (n = 616) | CKDu (n = 463) | P-value |
| --- | --- | --- | --- | --- |
| Age, mean (SD) (years) |  | 45.58 ± 8.21 | 52.07 ± 9.06 | **0.022** |
| Sex, n (%) | Male | 378 (61.36) | 96 (20.73) | **<0.001** |
|  | Female | 328 (38.64) | 367 (79.27) |  |
| Marital status, n (%) | Single | 46 (7.47) | 92 (19.87) | **<0.001** |
|  | Married | 570 (92.53) | 371 (80.13) |  |
| Physical activity, n (%) | 1<sup>st</sup>tertile (<=35.3) | 209 (33.93) | 151 (32.61) | **0.025** |
|  | 2<sup>nd</sup> Tertile (35.4-38.9) | 186 (30.19) | 174 (37.58) |  |
|  | 3<sup>rd</sup>tertile (>=39) | 221 (35.88) | 138 (29.81) |  |
| Low HDL n (%) | No | 319 (51.79) | 221 (47.73) | **0.187** |
|  | Yes | 297 (48.21) | 242 (52.27) |  |
| Hyper TG n (%) | No | 458 (74.35) | 290 (62.63) | **<0.001** |
|  | Yes | 158 (25.65) | 173 (37.37) |  |
| BMI n (%) | <18.5 | 68 (11.07) | 10 (2.16) | **<0.001** |
|  | 18.5-25 | 257 (41.86) | 140 (30.24) |  |
|  | >=25 | 289 (47.07) | 313 (67.60) |  |
| Smoking status n (%) | Non smoker | 446 (72.40) | 412 (88.98) | **<0.001** |
|  | Current smoker | 64 (10.39) | 19 (4.10) |  |
|  | Previous smoker | 106 (17.21) | 32 (6.91) |  |
| House use of pesticides n (%) | No | 365 (59.25) | 230 (49.68) | **0.002** |
|  | Yes | 251 (40.75) | 233 (50.32) |  |
| House use of pesticides exposure time (minutes) n (%) | Non users | 365 (59.25) | 230 (49.68) | **0.011** |
|  | 1<sup>st</sup> Tertile (<9) | 83 (13.47) | 82 (17.71) |  |
|  | 2<sup>nd</sup> Tertile (9-20) | 90 (14.61) | 72 (15.55) |  |
|  | 3<sup>rd</sup> Tertile (>20) | 78 (12.66) | 79 (17.06) |  |

In univariate analysis, female gender had the greatest crude odds ratio for CKDu (6.07, 95% CI 4.60-8.01). The effect of gender was raised in both multivariate models, compared to univariate analysis, reaching an odd of 7.52 (95% CI: 5.69-12.77) in model 1 and 8.64 (95% CI: 5.76-12.98) in model 2 (Table 2). Notably, the single marital status that had the second greatest odds ratio following female gender in univariate analysis (OR: 3.07, 95% CI: 2.11-4.48), was not associated with CKDu in multivariate analysis. Our analysis showed that age, hyper TG and BMI were among determinants of CKDu that kept their association in both models.

**Table 2.**
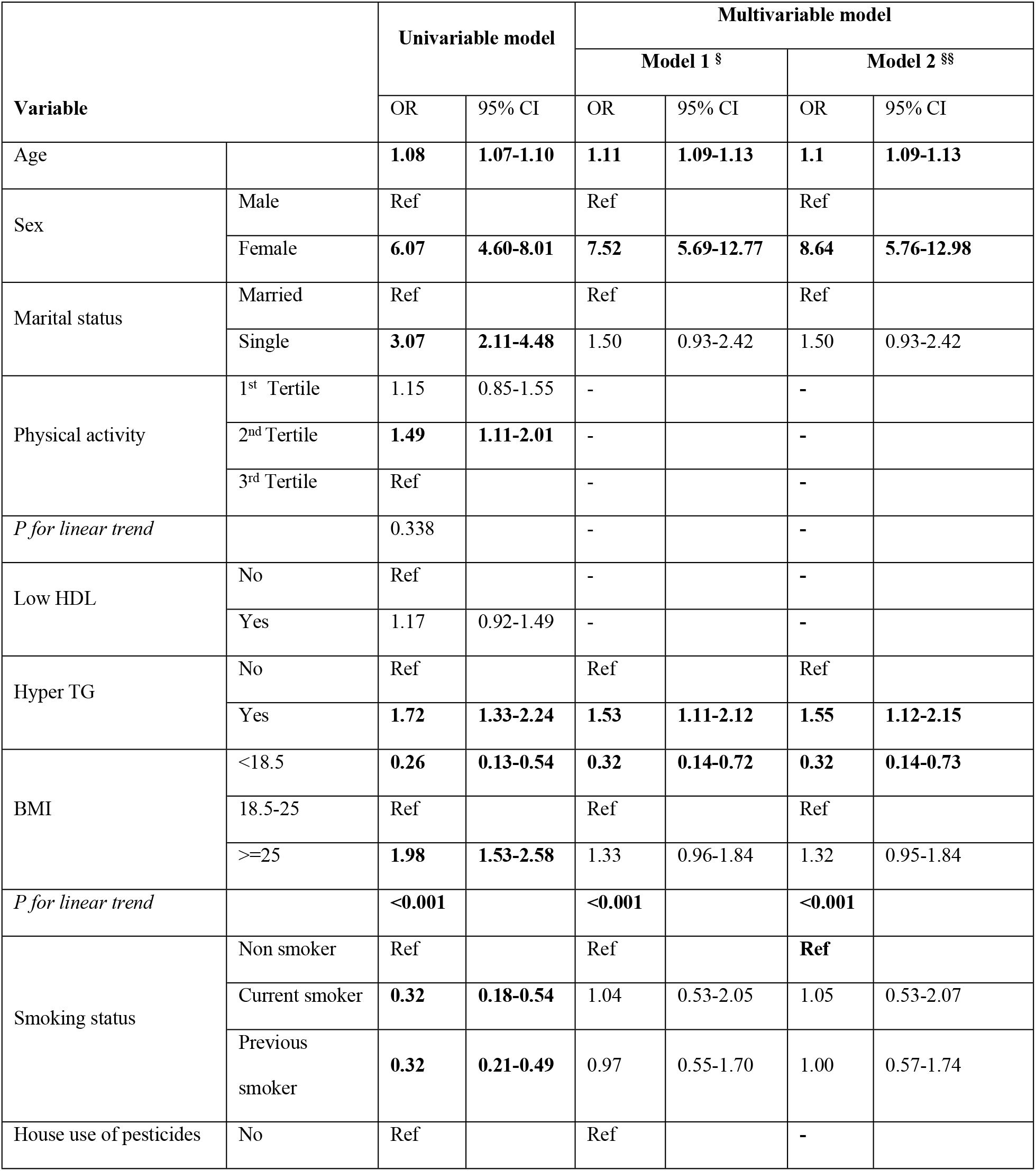
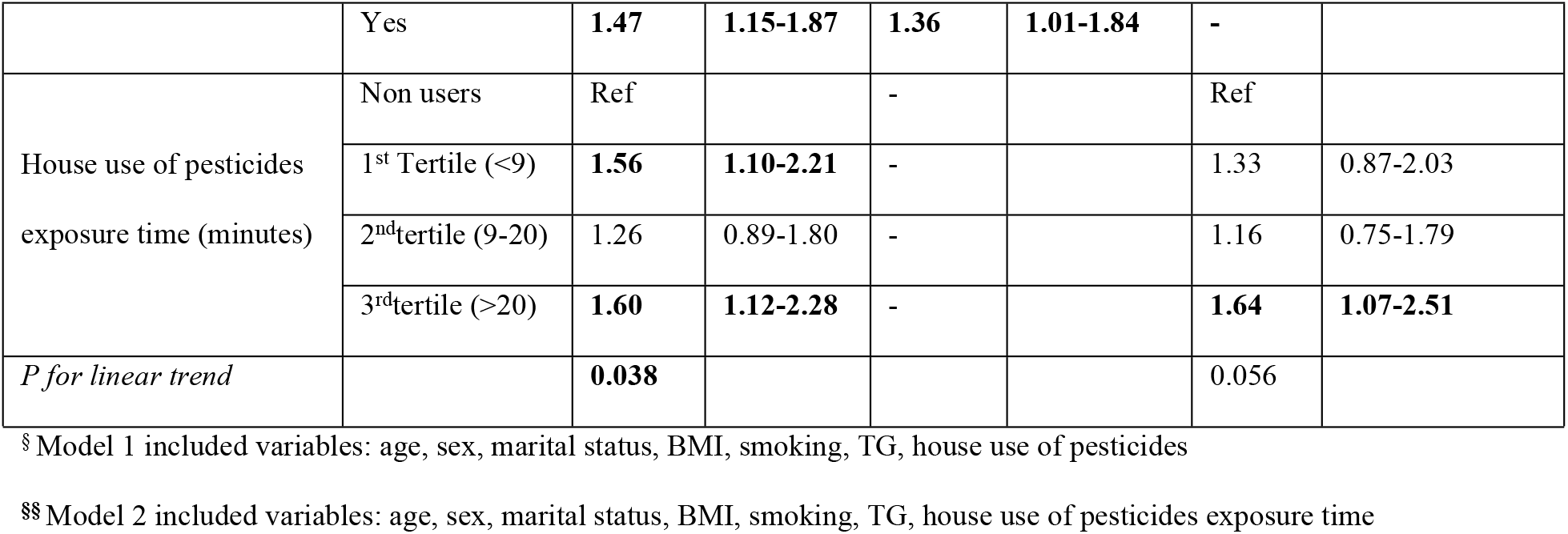
Risk factors for CKD of unknown origin in Univariable and Multivariable logistics regression analysis.

The house use of pesticides showed a significant association with CKDu in univariable analysis with an odds ratio of 1.47 (95% CI: 1.15-1.87); the effect was present in model 1. Duration of exposure to house pesticides had a significant association in univariable analysis; however, only the highest exposure time (3^rd^ tertile) had a significant association in model 2, in a way that subjects in this category showed 64% higher odds of CKDu than non-users.

## Discussion

This is the first study to investigate the determinants of CKDu with specific stress on household use of pesticides using data obtained through a population-based study in Iran.

The prevalence of CKD in this region in our study was 9.1% (95% CI 8.6-9.6) based on the MDRD equation. This figure was slightly lower than the findings of Tehran lipid glucose (TLGS)(24). The main reason might be the higher percentage of male subjects included in TLGS study, as many studies showed the higher prevalence of CKD among women compared to men(23,25–29). In another study in southwest of Iran, in Khuzestan region, the prevalence of CKD was reported slightly lower than Zahedan, at 7.1%(23). Younger age spectrum in Khuzestan study might at least partially explain the difference in CKD prevalence between two studies. The CKD prevalence in Golestan cohort study (GCS) on subjects older than 40 years old was reported at 23.7% (26.6% in women and 20.6% in men), which was twice our finding(25). While we cannot completely explain this variation, but difference in ethnicities could be among the reasons considering that more than three-quarters of the GCS population were Turkmen, while Sistan and Baluchestan population comprise of different ethnicities.

We found that in our study, prevalence of CKD in females was 2.6 times higher than male subjects, a proportion that is higher than other areas of our country and even other countries(23,25–29). For instance, a systematic review of CKD epidemiological features in Iran reported that prevalence of CKD in females was 1.7 times higher than in males, a figure that was much lower than our findings(3). The odds of having CKDu rises to 7.5 and 8.6 times in females when we analyzed in model including house use of pesticides (model 1) and pesticides exposure time (model2), respectively. This finding might point to risk of pesticides use as women spent more time at home in this area and subsequently, they might have an increased risk of pesticide exposure.

In our study, the time of house use of pesticides was significantly higher in subjects with CKDu than non-CKDu (50.3% vs. 40.8%), and the difference remained significant including in the multivariable model 1. Participants with a history of house use of pesticides have 1.36 times higher odds of CKDu than their counterparts. In Iran, low doses of organophosphates have been used in many insecticides, and handmade insecticides and pesticides are not uncommon in low-income families. Trabanino and colleagues described CKD of unknown origin for the first time in 2002; they found out most of these patients were farmers who had been exposed to pesticides regularly(30). In 2014, after many studies observed the impairment of kidney function among farmers, a study was designed on US Agricultural Health Study to determine the exposure-response trends among licensed pesticide applicators. They reported positive exposure trends in the use of herbicides alachlor, atrazine, metolachlor, paraquat, pendimethalin, and the insecticide permethrin in subjects with ESRD(31). Farmer’s wives were at risk of pesticides, which suggests that small amounts of pesticides and indirect exposure can also be harmful (31). Jayatilake et al. Found that the chronic low dose of cadmium exposure can result in CKD of unknown origin, and co-exposure to arsenic has an additive effect on impairment of kidney function(32). However, in a meta-analysis in 2018 in Meso-America, there was no association between pesticides exposure and GFR <60 ml/min/1.73 m^2^ as well (33). Since the chemical base of insecticides and pesticides are similar and pesticides may be used at home too, there is an urge to clear the effect of house use of pesticides and insecticides on kidney function.

We found a positive trend of exposure time of house use of pesticides on CKDu. Although the effect was faded in multivariable regression, the participants with exposure time in the highest tertile have 1.64 more odds of having CKDu than never users. This finding emphasizes the role of cumulative exposure dose at a specific time on kidney function. Although we cannot comment on safe threshold dose of house use of pesticides, as this was not in our study scope, but finding the safe use threshold of these materials could be of great interest that could be evaluated in longitudinal studies.

The prevalence of CKD without hypertension and diabetes mellitus was approximately 36% in our study, while it was much lower in studies in Malawi (0.2%) and India (1.6%). Even though participants with proteinuria were excluded from the mentioned studies, our finding was more than expected(34,35).

A low HDL did not have an association with CKDu in this study. Low level of HDL is commonly seen in the end stages of CKD due to impairment of reverse cholesterol transport(36). We should emphasis that all our subjects in CKDu group had CKD stage 3. This might explain why we could not see a significant effect of low HDL level on CKDu. Ghosh and colleagues demonstrated positive association of total lipid level with CKDu(37). The total lipid was calculated by sum of 2.27 times of total cholesterol, triglycerides plus 0.623 and did not address HDL and triglycerides separately. We also found that higher TG levels have a significant relation with CKDu, an effect that remained even in multivariable regression. The relation between dyslipidemia and CKD has been proposed by many studies(28,38–41). Studies showed that higher level of TG is seen in the early stages of CKD(42). A multicenter cohort study of non-dialysis dependent outpatients with CKD in Japan reported a decreasing effect of high TG with CKD staging as well (31). Taken together, this might justify the significant odds of high TG levels on CKDu, as all the participants categorized in CKDu had CKD stage3.

One limitation of this study is that since this study is part of the national cohort study, urine sampling and kidney sonography was not cost beneficial and were not performed. Thus, not all the CKDu patients had unknown etiology. However, we tried to minimize the known CKD patients with accurate screening for diabetes mellitus and HTN based on the PERSIAN cohort protocol. Another limitation of this study was a one-time measurement of serum creatinine, which can be misleading because of variation due to weather, season, and work pattern; Thus, our finding must be confirmed by future studies with at least two times measurement of creatinine. Although epidemiologic studies have internal limitations in assessing CKD and its etiologies, they are the first step in assessing risk factors and protective factors and should not be overlooked. Longitudinal studies should be carried out, and all subjects with the known etiologies of CKD (diabetes mellitus, systemic hypertension, primary glomerular nephritis, or structural abnormality) have to be excluded. Using novel biomarkers like KIM-1 and NGAL can facilitate the estimation of kidney damage in national epidemiologic studies as well(43,44).

## Conclusion

In this study, for the first time we assessed the prevalence of CKDu and possible determinants in a specific region in our country. We obtained our data through a population-based study. To our knowledge this is the first study of this kind that has been performed in our region. We found that indoor use of pesticides could be a possible risk factor for CKDu. Based on the previous research, pesticides can be potentially harmful in outdoor uses for kidney function. Our findings raise the concern of indoor use of pesticides on kidney function on subjects even without known risk factors of CKD including hypertension and diabetes. Further longitudinal studies should be carried out to assess the effect of indoor pesticides on CKD And to find the safe dose of these materials.

## Data Availability

Data cannot be shared publicly because of sensitive information. Data are available from the TUMS In Ethics Committee (contact via email of corresponding author) for researchers who meet the criteria for access to confidential data.

## Acknowledgments

We acknowledge all personnel in Zahedan center, who contributed a significant amount of time in collecting data, transferring blood samples, and making comfortable situation for participants during their interview and examinations.

## Financial Disclosure Statement

This study was funded by Zahedan University of Medical Sciences (Grant number: 93-4-96451). The Iranian Ministry of Health and Medical Education has also contributed to the funding used in the PERSIAN Cohort through Grant no. 700/152 and 700/534.

## Competing interests

The authors declare no competing interests.

## Notes

### Competing Interest Statement

The authors have declared no competing interest.

### Author Declarations

This study was approved by the ethics committee Zahedan University of Medical Sciences as the main executive university with the approval number of IR.ZAUMS.REC.1401.160.

